# A randomized sham-controlled trial of transcranial and intranasal photobiomodulation in Japanese patients with mild cognitive impairment and mild dementia due to Alzhimer’s disease: a protocol

**DOI:** 10.1101/2023.12.07.23299692

**Authors:** Yuma Yokoi, Takuma Inagawa, Yuji Yamada, Makoto Matsui, Asumi Tomizawa, Takamasa Noda

## Abstract

**Introduction:** Photobiomodulation (PBM) is a novel strategy for cognitive enhancement by improving brain metabolism and blood flow. It is potentially beneficial for patients with Alzheimer’s disease (AD). We present a study protocol for a randomised controlled trial designed to evaluate the efficacy and safety of PBM.

**Method and analysis:** This is a single-centre, parallel-group, randomised, sham-controlled study. We enrol patients with mild cognitive impairment or dementia due to AD and assigned them to receive either active or sham stimulation at home for 12 weeks, with three sessions per week (20 minutes each). The stimulation involves invisible near-infrared light delivered by five applicators (one in a nostril, one on the frontal scalp, and three on the occipital scalp). The primary outcome will be the mean change in the Alzheimer Disease Assessment Scale–cognition from baseline to Week 12. We will also measure cognitive function, activity of daily living, behavioural and psychological symptoms, and caregiver burden. We will collect data at clinics at baseline and Week 12 and remotely at home. We estimate a sample size of 30 (20 active and 10 sham) based on an expected mean difference of −6.9 and an SD of 4.8. We use linear models for the statistical analysis.

**Ethics and dissemination:** The National Center of Neurology and Psychiatry Clinical Research Review Board (CRB3200004) approved this study. The results of this study will be published in a scientific peer-reviewed journal.

Trial registration details Japan Registry of Clinical Trials jRCTs032230339.

**ARTICLE SUMMARY:** Strengths and limitations of this study

- This study aims to investigate the impact of PBM on the cognitive function and quality of life of patients diagnosed with mild cognitive impairment or dementia, using a well-defined sample population.
- A potential limitation of this study is that it is conducted at a single centre and does not adequately assess the long-term outcomes of PBM.

## INTRODUCTION

Dementia is a clinical syndrome characterized by impaired cognitive functions and reduced capacity to perform daily activities, as well as affecting the quality of life and social well-being of their caregivers. Prior to dementia, there is a stage of mild cognitive impairment, which is a condition of decreased cognitive performance that does not meet the criteria for dementia and preserves their ability for independent living(1). While some pharmacological treatments are available, such as cholinesterase inhibitors, memantine, and anti-amyloid antibodies, their efficacy and safety are still under debate. Anti-amyloid antibodies, in particular, have been recently approved in the United States and Japan, but their high cost, modest benefit, and potential risk of causing amyloid-related imaging abnormality, which can be fatal, raise questions about their clinical utility (2). Besides pharmacological interventions, several modifiable factors, such as physical activity, social engagement, and depression, have been identified as possible risk or protective factors for dementia (3).

Photobiomodulation (PBM) is a light therapy method that employs visible light (wavelengths around 400-700 nm) and near-infrared light (wavelengths around 810-1100 nm). Its potential was first unveiled in 1968 when Mester et al. reported the positive effects of PBM on hair growth and wound healing in mice using a ruby laser (4). Since then, PBM has been investigated under various terms, including low-reactive level laser therapy (LLLT). More recently, it has been shown that light-emitting diodes (LEDs), which are more cost-effective than lasers, can also produce comparable results. Consequently, PBM encompasses interventions that utilize various types of light sources beyond lasers.

The mechanism underlying PBM’s effects varies depending on the wavelength of the light used. Notably, light in the red-near-infrared region offers superior tissue penetration and contributes to cell proliferation and growth promotion(5)(6), Conversely, blue region light is expected to exhibit effects such as bactericidal effect and induction of cell apoptosis at high irradiation levels. In the case of low-level irradiation of red-near-infrared region light, it is believed that photons are absorbed by intracellular chromophores present in intracellular organelles (especially within mitochondria). Cytochrome C oxidase (CCO) is involved as a light receptor for this process(7), and the activation of CCO increases ATP production and activates cellular metabolic activity. It is also thought that the reactive oxygen species (ROS) and nitrogen oxide (NO) generated in this process promote gene transcription and cell repair. NO also has a role in regulating neurotransmitters in the central nervous system and vasodilation, which is thought to improve blood circulation. As a result, increased cerebral blood flow (8) and increased brain metabolism (9) have been observed with transcranial PBM.

The field of PBM has witnessed significant progress in improving brain stimulation parameters. Notably, the shift from continuous to pulsed stimulation has shown potential benefits, enhancing tissue absorption responses and possibly inducing specific brain wave rhythms, such as alpha waves at 10 Hz and gamma waves at 40 Hz (10). Furthermore, innovative techniques, like trans nasal olfactory nerve stimulation, have been developed to reach deeper brain regions (11). Coupled with the cost-effectiveness and miniaturization of LED devices, these improved parameters have heightened expectations for the effectiveness of PBM in neurological and psychiatric applications, especially in the context of cognitive enhancement. Consequently, a surge of clinical studies is expected to assess its impact in these domains. While the effectiveness of 40 Hz PBM on amyloid load in mice has yielded mixed results (12)(13), several clinical trials have shown potential in enhancing cognitive function in dementia patients.

For example, a sham-controlled clinical study by Chan et al. involved treating 18 patients with nine LEDs during a single stimulation session, which resulted in improved visual memory (14). Saltmarche et al. employed 5 patients pulsed 810 nm light at 10 Hz with an intensity of 14.2 mW/cm^2^ for sessions lasting 25 minutes, conducted once or twice weekly over 12 weeks, and observed improvements in Mini-Mental State Examination (MMSE (15)) and Alzheimer’s Disease Assessment Scale – Cognitive subscale (ADAS-Cog(16)) scores compared to baseline (17). Similarly,(18) Berman et al. recruited 11 patients and adopted pulsed 1072 nm light at 10 Hz for 6 minutes daily over 28 consecutive days, noting improvements in quantitative electroencephalogram compared to a sham stimulation group (19). Nizamutdinov et al. administered continuous 1060-1080 nm light at an intensity of 23.1 mW/cm^2^ for sessions lasting 6 minutes, twice daily over 12 weeks to 60 patients, and reported improvements in MMSE and LMT-I scores compared to a sham stimulation group (20).

While the above paper used transcranial PBM, another clinical study has also used a nasal applicator to stimulate the deep brain trans nasally. Chao et al. utilized pulsed 810 nm light at 40 Hz with an intensity of 75 mW/cm^2^ for 20-minute sessions thrice weekly over 12 weeks to 8 patients, reporting enhancements in ADAS-Cog and NPI scores compared to a sham stimulation group (18).This article also suggests increased cerebral perfusion and connectivity between the posterior cingulate cortex and lateral parietal nodes within the default-mode network in the PBM group. Our study try to extend this evidence with more number of participants in Japan to evaluate the utility of transcranial and trans nasal PBM in early Alzheimer’s disease (AD) patients, who are expected to exhibit mild neurodegeneration due to dementia.

## METHODS AND ANALYSIS

### Objective

The main objective of this study is to assess the effectiveness and safety of 12-week photobiomodulation on cognition among mild cognitive impairment and mild dementia due to AD patients as assessed by ADAS-Cog.

### Trial design

This is a prospective, parallel, randomised, sham-controlled, superiority trial involving 30 participants with AD-related mild cognitive impairment or dementia, as per the DSM-5 criteria. The participants will be allocated to either an active or a sham group in a 2:1 ratio. The protocol adheres to the 2013 SPIRIT guidelines. (21) The trial is registered with the Japan Registry of Clinical Trials (Trial ID: jRCTs032230339).

### Participants

We will enrol outpatients with regular consultations from a single academic hospital in Tokyo, Japan: the National Center of Neurology and Psychiatry. Potential participants will be referred by their treating psychiatrists or neurologists and receive a brochure with brief information on the trial after their initial appointments. Participants who express interest will provide informed consent to the principal investigator or subinvestigators using the Informed Consent Form (online supplemental file 2). The study design, benefits and risks will be explained to the participants by the principal investigator and outcome assessors. Participants will undergo screening by a treating psychiatrist to verify their eligibility (shown below). The inclusion criteria are: 1) Age between 60 and 90 years at consent, 2) Prior visit to the study site, 3) Early AD diagnosis (mild AD or MCI due to AD) confirmed by the study site, 4) MMSE score between 15 and 30 points, 5) Availability of a study partner who cohabits or regularly monitors the participant’s condition, 6) Ability to communicate and complete questionnaires via the Internet, email, etc. using a smartphone or personal computer (with or without study partner assistance), 7) Stable treatment for early AD (no change in drug or non-drug therapies for more than 4 weeks), 8) No major environmental changes expected within 12 weeks after the trial initiation, such as hospitalization, travel, relocation, etc. The exclusion criteria are as follows; 1) People with cognitive impairment from causes other than AD (such as mental disorders, intellectual disabilities, etc.), 2) People who join a new clinical trial or study during this study period, 3) People who are not suitable for this study according to the principal investigator or subinvestigators.

### Intervention

The study will employ a home-based PBM intervention using the Vielight Neuro Gamma 4 device (Vielight Inc., Tronto, Ontario, Canada), which delivers near-infrared emissions via five applicators (one intranasal, one frontal, and three occipital). The device will target the frontal, temporal, occipitoparietal, and naso-temporal regions. Subjects will blindly receive either active or sham devices from the investigator at the time of study entry. At that time, the investigator will adjust the device to the appropriate angle and instruct the subject and study partner on the appropriate way of its use. The subject and study partner will be responsible for the daily removal and recharging of the device, and they will have free access to the device instructions and instructional videos created for the study. They will also report and communicate regularly with researchers to check for malfunctions and failure of the device.

The participants will be randomised into two groups: active or sham PBM. The active group will receive 40-Hz intermittent photoemission per session with 60 J/cm2 on the frontal region, 45 J/cm2 on the occipital region in total, and 15 J/cm2 intranasally. The sham group will receive a placebo device that mimics the appearance of the active device but does not emit any light for 20 minutes per session. Both devices will have a green light indicator and will turn off automatically after 20 minutes. The participants and their study partners will be unable to discern whether they are using the active or sham device. The assessors and participants will be blinded to the treatment allocation to enhance the study validity. At the end of Week 12, the participants will be asked to guess their treatment group to assess the blinding quality. The participants and their study partners will also monitor and report any adverse events to the researchers remotely, as well as submit periodic reports of efficacy, safety, and satisfaction. The study partners will keep track of the device usage frequency and occasionally contact the participants or their partners to encourage adherence to the study protocol.

Any changes in the concomitant therapy of the participants, such as starting, stopping, or adjusting the dose, should be documented, and reported to the researcher. The researcher will train the patient and study partner on operating the device at home and provide written instructions before the first use. The researcher will also verify that the patient and study partner can use the device correctly. A video tutorial is available for reference at any time, and any issues or questions regarding the device use are regularly monitored to enhance adherence. The participant will be withdrawn from the study if any of the following conditions apply, as determined by the principal investigator or subinvestigators when the participant revokes their consent or asks to stop participating in the research; the participant is affected by a disease outbreak or a pandemic; the device shows insufficient efficacy for the participant; the participant fails to attend the evaluation and cannot be reached; the principal investigator or subinvestigators judge that the participant’s mental and physical burden is too high to continue in this study; there is a serious breach or violation of the research protocol. The entire study may be terminated by the Independent Safety and Effectiveness Committee, which convenes when three or more serious adverse events related to the study device occur.

## OUTCOMES

The main outcome of interest is mean change in ADAS-Cog scores from baseline to Week 12 between groups. Other outcomes include global function, assessed by Clinical Dementia Rating scale; quality of life, evaluated by the DEMQOL(22) and DEMQOL-proxy(23); cognition, measured by the MMSE (with a score range of 15 to 30); global impression, rated by Participant’s Global Impression of Improvement (PGI-I) and Clinical Global Impression of Improvement (CGI-I) ; behavioural and psychological symptoms, monitored by Neuropsychiatric Inventory Questionnaire (NPI(24)) ; and caregivers’ burden, measured by Zarit’s Burden Interview (ZBI(25)). A specialized psychologist, who was blind to group assignment, scored all these outcomes except DEMQOL, DEMQOL-proxy, PGI-I, NPI and ZBI after a clinical interview. The psychologists conducted the assessments at baseline, Week 12, and the follow-up. The psychological evaluations at Week 12 and the follow-up had a two-week window respectively. Figure 1 summarises the schedule for enrolment, intervention, and assessment. We will record any history of substance use disorder, dementia, psychiatric disorders, neurological disorders, brain injury and other conditions at baseline to exclude patients with clinically contraindicated conditions. Adverse events are any undesirable patient experience during PBM administration. We will monitor and record all adverse events throughout the study via self-report or clinical observations, regardless of causal relationship. We will follow up any untreatable adverse events after trial completion. The principal investigator will diagnose, treat, and explain any serious adverse events to the relevant patients. The subinvestigators will also report any information related to such adverse events to the principal investigator. The principal investigator is responsible for reporting any serious adverse events to the relevant authorities, including the clinical research review board, the Ministry of Health, Labour, and Welfare, and the pharmaceutical and medical device agencies.

**Figure 1.**
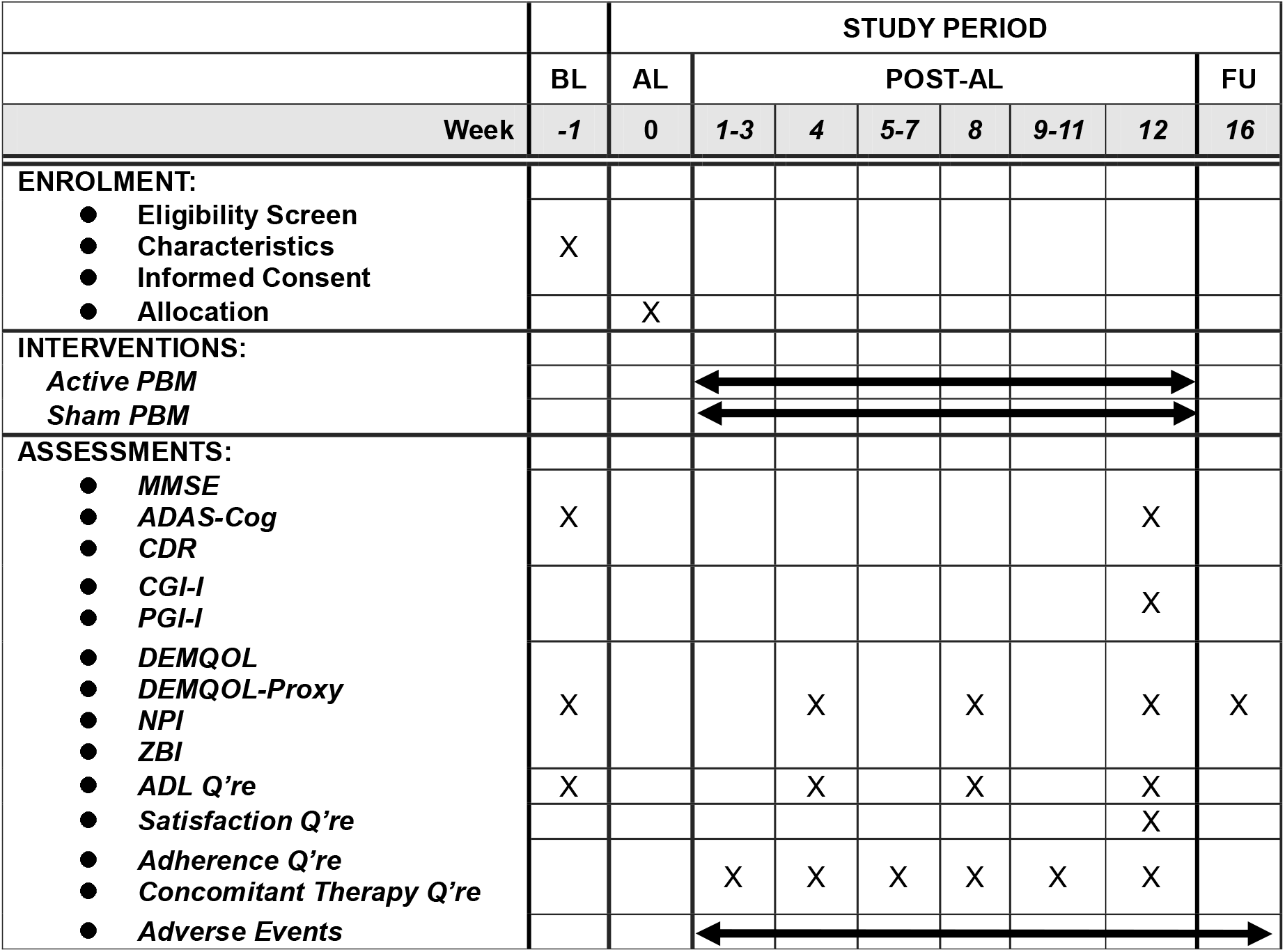
The schedule of this study The time point of post-tDCS evaluation will be allowed within ±7 days and that of follow-up evaluation will be allowed within 14 days. ADAS-Cog: Alzheimer’s Disease Assessment Scale – Cognitive Subscale, ADL: activities of daily living, AL: allocation, BL: baseline, CDR: Clinical Dementia Rating, CGI-I: Clinical Global Impression – Improvement, FU: follow-up, MMSE: Mini-Mental State Exam, NPI: Neuropsychiatric Inventory, PBM: photobiomodulation, Q’re: questionnaire, ZBI: Zarit Burden Interview

Serious adverse events are any undesirable medical occurrences at any dose that: 1. Cause death. 2. Pose a threat to life. 3. Necessitate inpatient hospitalisation or prolong existing hospitalisation. 4. Lead to persistent or significant disability/incapacity. 5. Are congenital anomalies/birth defects. 6. Require interventions to prevent permanent impairment damage. The psychiatrists involved in the treatment will document the name, occurrence date, severity, grade, intervention implemented (if any), outcome and relationship to the study medication in an electronic clinical research form (eCRF). Any symptoms that were present at baseline and did not worsen during the study will not be considered as adverse events. Adverse events are classified into three categories in this study: Mild: asymptomatic or mild symptoms; clinical or diagnostic observations only; no intervention required; no impact on usual daily activities. Moderate: minimal, local or non-invasive intervention required; limiting age-appropriate instrumental activities of daily living (ADL); some impact on usual daily activities. Severe: medically significant, life-threatening or related to death; hospitalisation or the prolongation of hospitalisation required; disabling; limiting self-care in ADL; significant impact on usual daily activities.

### Sample size calculation

We aim to enroll 30 participants. In Chao et al.’s report (18), the intervention group scored 32.3±4.8 and the control group scored 39.2±2.6 on ADAS-Cog, which is an efficacy evaluation item for cognitive function. In this trial, assuming a 1:2 allocation ratio of sham stimulation group to real stimulation group, an ADAS-Cog score of 39.2 for the sham stimulation group, an ADAS-Cog score of 32.3 for the real stimulation group, a standard deviation of 4.8, α=0.05, and a power of 0.80, the required sample size was calculated to be 18 (6 for the sham stimulation group and 12 for the real stimulation group) using https://clincalc.com/stats/samplesize.aspx. Considering that this study includes patients with mild cognitive impairment, which is a preliminary stage of the previous study, and that the ADAS-Cog score may decrease overall, we multiplied the sample size by 1.5 to obtain 27, and further considered adherence and dropout cases to obtain 30.

### Sequence generation

We will use a computer-generated stratified sequence from the Mujinwari system (https://mujinwari.biz/) to assign participants to the active or sham PBM group in a 2:1 ratio. Our custom electronic data capture (EDC) system, based on Google Workspace For Business (Google, USA), will collect all data. A cloud computer will keep group assignment and other personal identification data. It will ensure a balanced distribution of diagnosis types (e.g., major neurocognitive disorder or mild neurocognitive disorder). This randomisation method stratifies by diagnosis, and the independent personnel can access the allocation results and provide the appropriate device. The principal investigator and subinvestigators can access each participant’s allocation and disclose it to the participants and raters only after the complete study data is consolidated.

### Data collection methods and data management

To evaluate the intervention, we will perform assessments by clinicians at baseline and Week 12 (figure 1). Experienced psychologists unaware of the group allocations will conduct the baseline, Week 12 and follow-up evaluations. Participants and study partners will complete scheduled questionnaires and report adverse events between these visits. All data, except for the informed consent document, will be entered into the EDC anonymously and verified by independent data managers. Considering the device’s safety and the blindness of study personnel, we will not composite the specific data monitoring committee, but data monitors will oversee and review the trial progress. Participants can withdraw their consent or discontinue participating in the study at any time, but we will collect data as much as possible unless consent is withdrawn, or it is at least a serious deviation from the protocol. We will terminate the intervention if we observe any serious adverse events (SAEs). The following auditing standards for field work apply to this study based on the Clinical Research Act: audits will be conducted if any of the following criteria are met: (1) at least two SAEs that are causally related to the medical device are reported; (2) at least two serious protocol deviations are detected; (3) multicentre clinical trials are initiated; or (4) potential severe conflicts of interest that deviate from the prespecified plan for conflicts of interest are identified. We will report any necessary protocol amendments and their outcomes to both the clinical research review board and the Ministry of Health, Labour, and Welfare for registration on the Japan Registry of clinical trials website. After completing the trial, we will publish an original article to disseminate the data results.

### Patient and public involvement

None.

### Statistical analysis

We define two populations for our analyses. Full Analysis Set (FAS) will be those participants who are randomised and observed for efficacy assessment after taking active or sham devices home; Safety Analysis Set will be those who are randomised and observed for any safety assessment. Our primary analysis for efficacy will be FAS. Our primary analysis will be intention-to-treat and include all randomised patients in the active or sham group, and we will report their demographic characteristics. We will also perform per-protocol set analysis after excluding cases with protocol deviations as sensitivity analyses. To estimate the mean treatment effect, we will use a linear model for analysis of covariance (ANCOVA) to detect changes in ADAS-Cog, MMSE, CDR, PGI-I, CGI-I, DEMQOL, DEMQOL-Proxy, NPI, and ZBI as secondary outcomes at baseline and Week 12. The ANCOVA will incorporate the covariates of group, baseline score of each outcome and disease (mild cognitive impairment or dementia), which are the stratification factors of dynamic allocation. We will not plan any interim analyses. The primary statistical test will be a t-test for the difference of the adjusted means of ADAS-Cog between the groups at Week 12. We will also conduct t-tests for the follow-up period and/or the other outcomes. For missing data, we will impute the baseline score as a sensitivity analysis. We will use a Fisher’s exact test to assess the blinding integrity. Additionally, we will describe the demographic characteristics of the patients and compare them between the two groups using a two-sample t-test or Fisher’s exact test. We will also examine the correlations between the baseline characteristics, ADAS-Cog, MMSE, CDR, DEMQOL, DEMQOL-proxy, NPI-Q, and ZBI and the cognitive outcomes at Week 12 and follow-up. We will use SAS V.9.4 or later for the statistical analysis. The results will be significant at p<0.05 and statistical tests will be conducted for two-tailed hypotheses.

## DISCUSSION

Since dementia is a condition that involves insidious deterioration of cognitive and social functioning, the long-term effects of certain intervention should be clinically important. Thus, we should conduct a long-term trial with minimizing number of visits in hospitals to enhance adherence to the intervention. To this end, this device is suitable for home use, convenient and proper to wear or remove it. Besides applications at home needs fundamental safety of the device. This device has gained the status of General Wellness Device from the US Food and Drug Administration due to the safety and ability of home use. The remaining problem is the resource to conduct a huge clinical trial with long-term duration. Most other studies cannot overcome this and PBM studies have limited sample size generally. This study also fell to prey but the positive results of the properly conducted study hopefully evoke further and bigger clinical studies.

### Strengths and limitations of this study

This study has several strengths, such as its rigorous sample size estimation for cognitive outcomes based on prior evidence, in contrast to many previous studies that did not perform adequate power calculations. Moreover, the device used in this study is unique in its ability to deliver near-infrared light through the nasal cavity. Given the long duration of 12 weeks for the participants and their caregivers, we implemented strategies to enhance adherence, such as minimizing the number of hospital visits and providing immediate contact with the researchers. The use of remote visits also improved the experience of the researchers. However, we also acknowledge some limitations of this study. First, the study is conducted at a single site, which may affect the generalizability of the findings. Second, the long-term effects of PBM on cognition cannot be evaluated. This is a common challenge in trials of anti-amyloid therapies, which require 78 weeks or more to show results and are still controversial regarding their sustained efficacy. Therefore, the optimal duration of PBM intervention remains an open question.

## ETHICS AND DISSEMINATION

We have submitted the protocol version 1.02 to an institutional review board for ethical approval by the National Center of Neurology and Psychiatry Clinical Research Review Board (CRB3200004), in accordance with the Decla ration of Helsinki and the Ethical Guidelines for Medical and Health Research Involving Human Subjects. The institutional clinical research review board will conduct initial and annual reviews of the protocol, starting from 1 September 2023. The principal investigator will provide safety and progress reports to the review board at least once a year, and within 3 months after the study termination or completion. These reports will contain the total number of patients enrolled, serious and non-serious adverse events that occurred, and summaries of the safety and monitoring board’s review. The clinical research review board and the Ministry of Health, Labour, and Welfare will receive prompt notification of any SAEs from the principal investigator. This study complies with the Clinical Trials Act and is registered with the Japan Registry of Clinical Trials. The study objectives will be explained to all study participants, who will provide informed consent before the principal investigator, the research coordinator or the research assistant enrols them in the trial. Participation is voluntary and patients will undergo assessment after consenting. The clinical research review board will approve any protocol amendments that involve changes in the study protocol and/or the informed consent. Participants who need medical treatment will access it through their own medical insurance.

Regarding data access and responsibility, the designated investigators (YY and TI) has full access to all of the data in the study and takes responsibility for the integrity of the data and the accuracy of the analysis. Individual participant data generated in this study will be anonymised for transparency of trial results and adherence to ICMJE recommendations.

They will be shared with researchers for secondary use of the data upon request per the data-sharing policy. The data curation will be completed by January 2026 and the results will be published by March 2026. The data will be retained for 5 years after the study completion.

## Data Availability

All data produced in the present study are available upon reasonable request to the authors

## Data Sharing

Data sharing:Technical appendix, statistical code, and dataset available from the corresponding author at.

## Author Contributions

Yuma Yokoi developed the original concept for the trial. MO advised a statistical analysis plan for the original protocol, and YY, TI and MO established the protocol. YY and TI will recruit the participants. MM will evaluate the participants. TN will supervise the whole progress of this research. YY wrote the manuscript, and all other authors reviewed and commented on the subsequent drafts. All authors read and approved the final manuscript.

## Acknowledgements

We would like to express our gratitude to Dr. Mari Oba, who provided valuable statistical advice for this study.

## SPIRIT 2013 Checklist

| Section/item | Item No | Description | Addressed on page number |
| --- | --- | --- | --- |
| **Administrative information** |  |  |  |
| Title | 1 | Descriptive title identifying the study design, population, interventions, and, if applicable, trial acronym | p.1 |
| Trial registration | 2a | Trial identifier and registry name. If not yet registered, name of intended registry | p.2, p.3 |
|  | 2b | All items from the World Health Organization Trial Registration Data Set | p.2, p.3 |
| Protocol version | 3 | Date and version identifier | p.7 |
| Funding | 4 | Sources and types of financial, material, and other support | p.8 |
| Roles and responsibilities | 5a | Names, affiliations, and roles of protocol contributors | p.1 |
|  | 5b | Name and contact information for the trial sponsor | p.1 |
|  | 5c | Role of study sponsor and funders, if any, in study design; collection, management, analysis, and interpretation of data; writing of the report; and the decision to submit the report for publication, including whether they will have ultimate authority over any of these activities | p.8 |
|  | 5d | Composition, roles, and responsibilities of the coordinating centre, steering committee, endpoint adjudication committee, data management team, and other individuals or groups overseeing the trial, if applicable (see Item 21a for data monitoring committee) | p.7 |
| **Introduction** |  |  |  |
| Background and rationale | 6a | Description of research question and justification for undertaking the trial, including summary of relevant studies (published and unpublished) examining benefits and harms for each intervention | p.2 |
|  | 6b | Explanation for choice of comparators | p.3 |
| Objectives | 7 | Specific objectives or hypotheses | p.3 |
| Trial design | 8 | Description of trial design including type of trial (eg, parallel group, crossover, factorial, single group), allocation ratio, and framework (eg, superiority, equivalence, noninferiority, exploratory) | p.3 |
| **Methods: Participants, interventions, and outcomes** |  |  |  |
| Study setting | 9 | Description of study settings (eg, community clinic, academic hospital) and list of countries where data will be collected. Reference to where list of study sites can be obtained | p.4 |
| Eligibility criteria | 10 | Inclusion and exclusion criteria for participants. If applicable, eligibility criteria for study centres and individuals who will perform the interventions (eg, surgeons, psychotherapists) | p.4 |
| Interventions | 11a | Interventions for each group with sufficient detail to allow replication, including how and when they will be administered | p.4 |
|  | 11b | Criteria for discontinuing or modifying allocated interventions for a given trial participant (eg, drug dose change in response to harms, participant request, or improving/worsening disease) | p.4 |
|  | 11c | Strategies to improve adherence to intervention protocols, and any procedures for monitoring adherence (eg, drug tablet return, laboratory tests) | p.4 |
|  | 11d | Relevant concomitant care and interventions that are permitted or prohibited during the trial | p.4 |
| Outcomes | 12 | Primary, secondary, and other outcomes, including the specific measurement variable (eg, systolic blood pressure), analysis metric (eg, change from baseline, final value, time to event), method of aggregation (eg, median, proportion), and time point for each outcome. Explanation of the clinical relevance of chosen efficacy and harm outcomes is strongly recommended | p.5 |
| Participant timeline | 13 | Time schedule of enrolment, interventions (including any run-ins and washouts), assessments, and visits for participants. A schematic diagram is highly recommended (see Figure) | p.5 |
| Sample size | 14 | Estimated number of participants needed to achieve study objectives and how it was determined, including clinical and statistical assumptions supporting any sample size calculations | p.6 |
| Recruitment | 15 | Strategies for achieving adequate participant enrolment to reach target sample size | p.5 |
| **Methods: Assignment of interventions (for controlled trials)** |  |  |  |
| **Allocation:** |  |  |  |
| Sequence generation | 16a | Method of generating the allocation sequence (eg, computer-generated random numbers), and list of any factors for stratification. To reduce predictability of a random sequence, details of any planned restriction (eg, blocking) should be provided in a separate document that is unavailable to those who enrol participants or assign interventions | p.6 |
| Allocation concealment mechanism | 16b | Mechanism of implementing the allocation sequence (eg, central telephone; sequentially numbered, opaque, sealed envelopes), describing any steps to conceal the sequence until interventions are assigned | p.6 |
| Implementation | 16c | Who will generate the allocation sequence, who will enrol participants, and who will assign participants to interventions | p.6 |
| Blinding (masking) | 17a | Who will be blinded after assignment to interventions (eg, trial participants, care providers, outcome assessors, data analysts), and how | p.6 |
|  | 17b | If blinded, circumstances under which unblinding is permissible, and procedure for revealing a participant's allocated intervention during the trial | p.6 |
| **Methods: Data collection, management, and analysis** |  |  |  |
| Data collection methods | 18a | Plans for assessment and collection of outcome, baseline, and other trial data, including any related processes to promote data quality (eg, duplicate measurements, training of assessors) and a description of study instruments (eg, questionnaires, laboratory tests) along with their reliability and validity, if known. Reference to where data collection forms can be found, if not in the protocol | p.6 |
|  | 18b | Plans to promote participant retention and complete follow-up, including list of any outcome data to be collected for participants who discontinue or deviate from intervention protocols | p.6 |
| Data management | 19 | Plans for data entry, coding, security, and storage, including any related processes to promote data quality (eg, double data entry; range checks for data values). Reference to where details of data management procedures can be found, if not in the protocol | p.7 |
| Statistical methods | 20a | Statistical methods for analysing primary and secondary outcomes. Reference to where other details of the statistical analysis plan can be found, if not in the protocol | p.7 |
|  | 20b | Methods for any additional analyses (eg, subgroup and adjusted analyses) | p.7 |
|  | 20c | Definition of analysis population relating to protocol non-adherence (eg, as randomised analysis), and any statistical methods to handle missing data (eg, multiple imputation) | p.7 |
| **Methods: Monitoring** |  |  |  |
| Data monitoring | 21a | Composition of data monitoring committee (DMC); summary of its role and reporting structure; statement of whether it is independent from the sponsor and competing interests; and reference to where further details about its charter can be found, if not in the protocol. Alternatively, an explanation of why a DMC is not needed | p.6 |
|  | 21b | Description of any interim analyses and stopping guidelines, including who will have access to these interim results and make the final decision to terminate the trial | p.7 |
| Harms | 22 | Plans for collecting, assessing, reporting, and managing solicited and spontaneously reported adverse events and other unintended effects of trial interventions or trial conduct | p.6 |
| Auditing | 23 | Frequency and procedures for auditing trial conduct, if any, and whether the process will be independent from investigators and the sponsor | p.7 |
| **Ethics and dissemination** |  |  |  |
| Research ethics approval | 24 | Plans for seeking research ethics committee/institutional review board (REC/IRB) approval | p.8 |
| Protocol amendments | 25 | Plans for communicating important protocol modifications (eg, changes to eligibility criteria, outcomes, analyses) to relevant parties (eg, investigators, REC/IRBs, trial participants, trial registries, journals, regulators) | p.7 |
| Consent or assent | 26a | Who will obtain informed consent or assent from potential trial participants or authorised surrogates, and how (see Item 32) | p.4 |
|  | 26b | Additional consent provisions for collection and use of participant data and biological specimens in ancillary studies, if applicable | Not applicable |
| Confidentiality | 27 | How personal information about potential and enrolled participants will be collected, shared, and maintained in order to protect confidentiality before, during, and after the trial | p.6 |
| Declaration of interests | 28 | Financial and other competing interests for principal investigators for the overall trial and each study site | p.9 |
| Access to data | 29 | Statement of who will have access to the final trial dataset, and disclosure of contractual agreements that limit such access for investigators | p.9 |
| Ancillary and post-trial care | 30 | Provisions, if any, for ancillary and post-trial care, and for compensation to those who suffer harm from trial participation | p.9 |
| Dissemination policy | 31a | Plans for investigators and sponsor to communicate trial results to participants, healthcare professionals, the public, and other relevant groups (eg, via publication, reporting in results databases, or other data sharing arrangements), including any publication restrictions | p.9 |
|  | 31b | Authorship eligibility guidelines and any intended use of professional writers | p.8 |
|  | 31c | Plans, if any, for granting public access to the full protocol, participant-level dataset, and statistical code | p.9 |
| **Appendices** |  |  |  |
| Informed consent materials | 32 | Model consent form and other related documentation given to participants and authorised surrogates | Attached |
| Biological specimens | 33 | Plans for collection, laboratory evaluation, and storage of biological specimens for genetic or molecular analysis in the current trial and for future use in ancillary studies, if applicable | Not applicable |

